# Single-Session Feasibility of a Wearable Music-based Rhythmic Auditory Stimulation Device to Improve Walking in Knee Osteoarthritis

**DOI:** 10.1101/2024.09.20.24313422

**Authors:** Yiwen Yang, Michele Okamura, Ehyun Kim, Deepak Kumar

## Abstract

**Objective:** We examined the single-session feasibility of a wearable device to deliver a closed-loop music-based rhythmic auditory stimulation (RAS) walking intervention in people with knee osteoarthritis (OA).

**Methods:** Fifteen participants with knee OA (age 70.0 ± 8.6 years; BMI 29.4±5.9 kg/m^2^; 10 women) complete a supervised 30-minute walking session using a wearable music-based RAS device. The device included shoe-worn inertial sensors, headphones, and a proprietary algorithm embedded in a touchscreen control unit. The algorithm continuously assessed users’ gait and adjusted the tempo of the selected music to encourage improvements in gait quality. Spatiotemporal gait measures were collected using separate inertial sensors during the session. We examined change in gait during the session, participant’s report of exertion, and participant-reported acceptability.

**Results:** On average, participants showed ∼12%, 30%, and 15% increases in cadence, gait speed, and stride length, respectively without worsening variability or symmetry from before to during the RAS session. Of the 15 participants, 9/15, 11/15, and 9/15 showed ≥ 10% increase in cadence, gait speed, and stride length, respectively. Rating of Perceived Exertion was on average 13.3 (range 12-15). There were no serious adverse events and small increase in knee pain that resolved at the end of the session. Participants were highly accepting of the device and willing to use it at home for longer periods.

**Conclusion:** A single-session feasibility study of a wearable closed-loop music-based RAS device in people with knee OA demonstrated increases in cadence and gait speed while safely walking at a moderate exercise intensity under supervision.

## INTRODUCTION

People with knee osteoarthritis (**OA**) experience impairments in both quality and quantity of walking. In terms of quality, altered gait patterns, including low cadence, slow gait speed, and alterations of knee kinematics, kinetics, and muscle activation, are all well-known characteristics of knee OA (1–5). These gait alterations can worsen both structural and symptomatic course of knee OA (3, 6–8). Deficits in walking quantity are also ubiquitous with individuals with knee pain walking 28% fewer steps/day on average compared to people without knee pain and over 50% of men and nearly 80% of women with or at risk of knee OA not meeting physical activity guidelines (9). Given that regular physical activity is a core recommendation for managing knee pain, reductions in physical activity can worsen knee pain leading to a vicious cycle (10, 11). Hence, there is a need for innovative interventions that can address deficits in both quality and quantity of walking in people with knee OA. However, such interventions currently do not exist.

We have previously reported that walking with low cadence, i.e., taking fewer steps per minute, is related to increased risk of worsening pain and physical function in people with knee OA (12). Walking with higher cadence could not only improve walking quality but also evoke a metabolic response consistent with light-to-moderate intensity exercise (13). As such, strategies to increase walking cadence could address impairments in both quality and quantity of walking for people with knee OA. Rhythmic auditory stimulation (**RAS**) is an approach that has been successfully used to improve walking related disability in people with neurological conditions (14–18). RAS interventions provide an external auditory cue (e.g., metronome) and rely on auditory-motor entrainment to achieve an improvement in walking performance. Auditory-motor entrainment refers to an involuntary synchronization of motor and auditory rhythms due to the entrainment of the neurons in the motor cortex via direct connections between the auditory and motor regions of the brain (14, 19, 20). A Cochrane review reported that RAS improves cadence (10.8 steps/min), gait speed (0.09 m/s), and stride length (0.12 m) when compared to usual therapy or gait retraining, in people with stroke (15). In people with Parkinson’s disease, RAS has been reported to improve gait speed and stride length with variable effects on cadence (21, 22). RAS has also been used effectively in people with traumatic brain injury, multiple sclerosis, and cerebral palsy (23–25). However, RAS has not been examined as an intervention to increase walking cadence in people with knee OA.

While RAS is typically delivered using a metronome, using music as the auditory cue can further enhance the effects of RAS (26–33). Music itself has also been explored as an intervention for chronic pain; while the specific mechanisms by which music alleviates pain are not fully elucidated, music is thought to modulate the activity in the pain and emotion networks of the brain (34–37). Music also evokes hedonic responses (e.g. positive affect, reduced perceived exertion), which could lead to hedonic motivation (i.e. desire to move) and improve adherence, a key barrier for people with knee OA (38). Finally, using wearable technology to deliver music-based RAS in a closed loop could drive sustained benefits by adapting to an individual’s walking performance throughout each session and over the course of the intervention. Such a digital intervention may also facilitate self-efficacy and independence by allowing access to the intervention in the person’s everyday life, outside of clinical settings.

Our objective was to examine the feasibility of a novel wearable device to deliver a music-based closed-loop RAS intervention in people with knee OA. Specifically, we examined (a) whether people with knee OA can safely increase their cadence and speed while using the device, (b) whether walking with the device yields a light-to-moderate intensity exercise experience, and (c) whether the participants find the device acceptable.

## PARTICIPANTS AND METHODS

### Participants

We recruited the participants from a database of previous participants and online advertisements on Craigslist. Participants were included if they were ≥50 years, had activity related pain, experienced morning knee stiffness ≤30 minutes, had a BMI<40, experienced knee pain on most days for 3 months or more, were able to walk at least 20 minutes without assistance, had knee pain severity of ≥4 on a 11-point numeric rating scale during previous week, and were able to communicate using English at a level to understand the study procedures and informed consent. We excluded people with contraindications to exercise, neurological conditions (e.g., stroke, Parkinson’s disease, Alzheimer’s disease, Multiple Sclerosis, diabetic neuropathy, etc.), joint replacement in either hip or ankle, history of other diseases that may involve the index joint including inflammatory joint disease such as rheumatoid arthritis, seronegative spondyloarthropathy, crystalline disease, lupus erythematosus, knee joint infections, Paget’s disease affecting the knee, or knee joint tumors, previous knee osteotomy partial or total knee replacement in either knee, corticosteroid or hyaluronic acid injections in either knee in the previous 3 months, pregnancy, and suspected or known drugs or alcohol abuse. Interested participants completed an online pre-screening questionnaire, followed by a screening phone call with a researcher. Those eligible were then scheduled for a single study visit. During the visit, the index knee was then defined as the more painful knee or a knee selected at random if both knees were equally painful. All study procedures were approved by the Institutional Review Board and all participants signed an informed consent before any study procedures. This device feasibility study is reported in compliance with the Strengthening the Reporting of Observational Studies in Epidemiology (STROBE) guidelines for cross-sectional studies.

### Wearable music-based RAS device (**Figure 1**)

The device consists of a kit including a touchscreen device (Samsung, Korea) locked with a preloaded proprietary software application (MedRhythms, Portland, ME), a bone conducting headset for delivery of RAS (Shokz, Austin, TX), and two shoe-worn inertial sensors (Physilog, Switzerland). Bone conduction earphones allow the user to still be able to hear other sounds while music is playing for safety. In the walking session, after a brief baseline period without music, a 30-minute overground walking session with music commenced and ended with a brief carryover walking period. The user was guided through the full session via audio instructions. The user chose a music playlist from a select list of available genres. Participants were instructed to begin walking at their regular pace to calibrate the digital intervention during the baseline period without music, and record the participant’s baseline gait parameters for that session. This was followed by the 30-minute walking session where personalized audio cues were embedded into time-shifted music based on real-time decisions made by a closed-loop control algorithm that continuously assesses the gait data collected by the inertial sensors. The algorithm tracked the user’s ability to entrain to the target tempo and also assessed gait quality (i.e., symmetry, variability). Both entrainment and quality were used by the algorithm to tailor the intervention to a person’s gait for personalized intervention during each session. The algorithm also added a rhythm track overlay to the music if needed to further assist with entrainment by increasing the salience of the beat. Participants were allowed to take breaks as needed. A researcher accompanied the participant during the walk.

**FIGURE 1.**
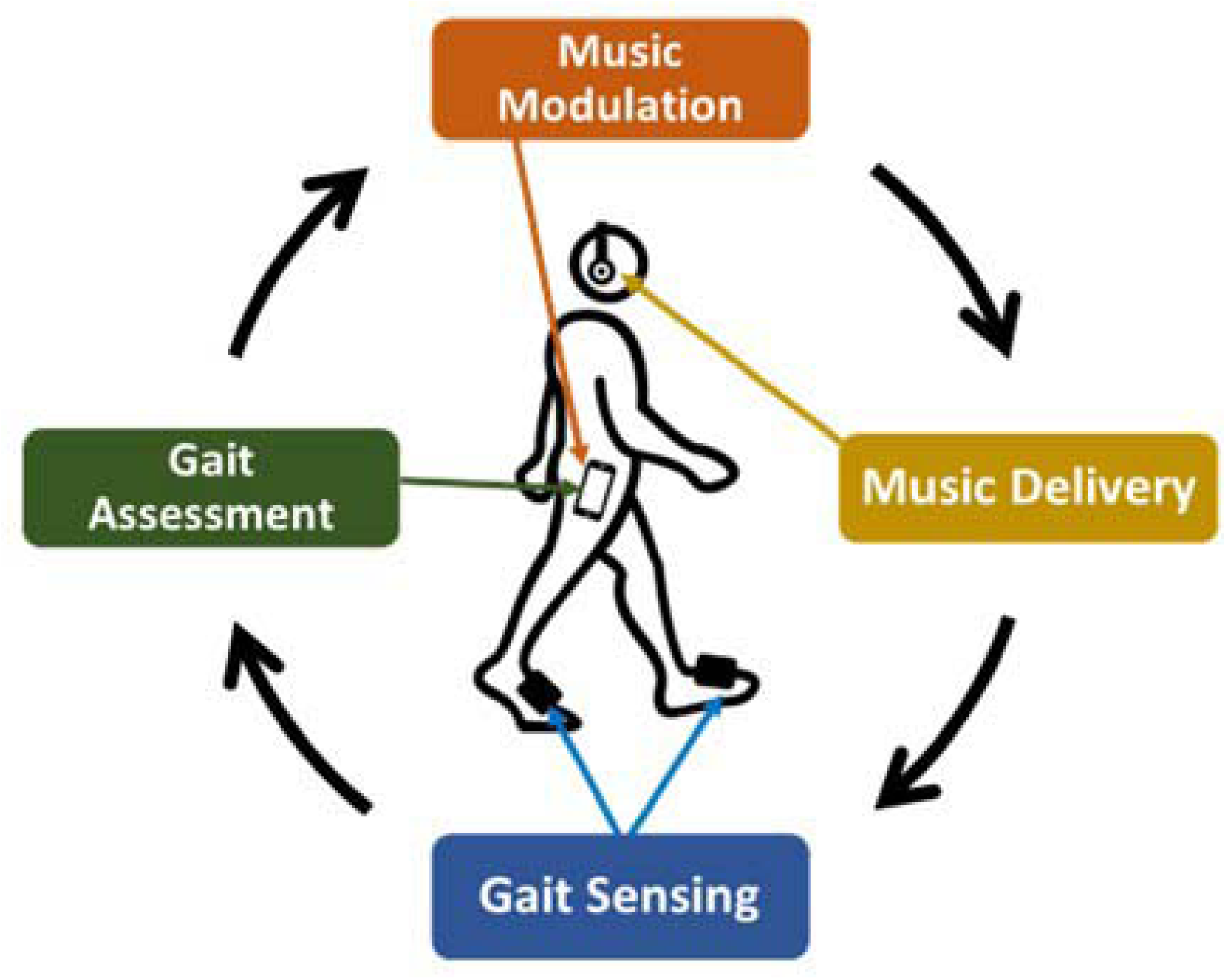
Wearable closed-loop music-based RAS intervention.

### Gait measures

In addition to the sensors worn as part of the RAS device, participants were instrumented with three additional inertial sensors (OPAL, APDM Inc, Portland, OR) worn on each shoe and the lower back, prior to the RAS session. Sensor data from these OPAL sensors were collected throughout the baseline, training, and carryover periods of the RAS session. Manufacturer-provided validated algorithms were used to extract spatiotemporal gait measures (e.g., cadence, gait speed, stride length, etc.) (39, 40). **Figure 2** shows cadence (measured by OPAL sensors) and instances when the tempo of the music was altered by the algorithm during the training part of the RAS session (excluding baseline and carryover) for a representative participant. We additionally calculated measures of gait variability and symmetry from the spatiotemporal measures. Variability was defined as the coefficient of variation (CoV) of step duration and symmetry between left and right step duration was calculated using the symmetry index method (41).

**FIGURE 2.**
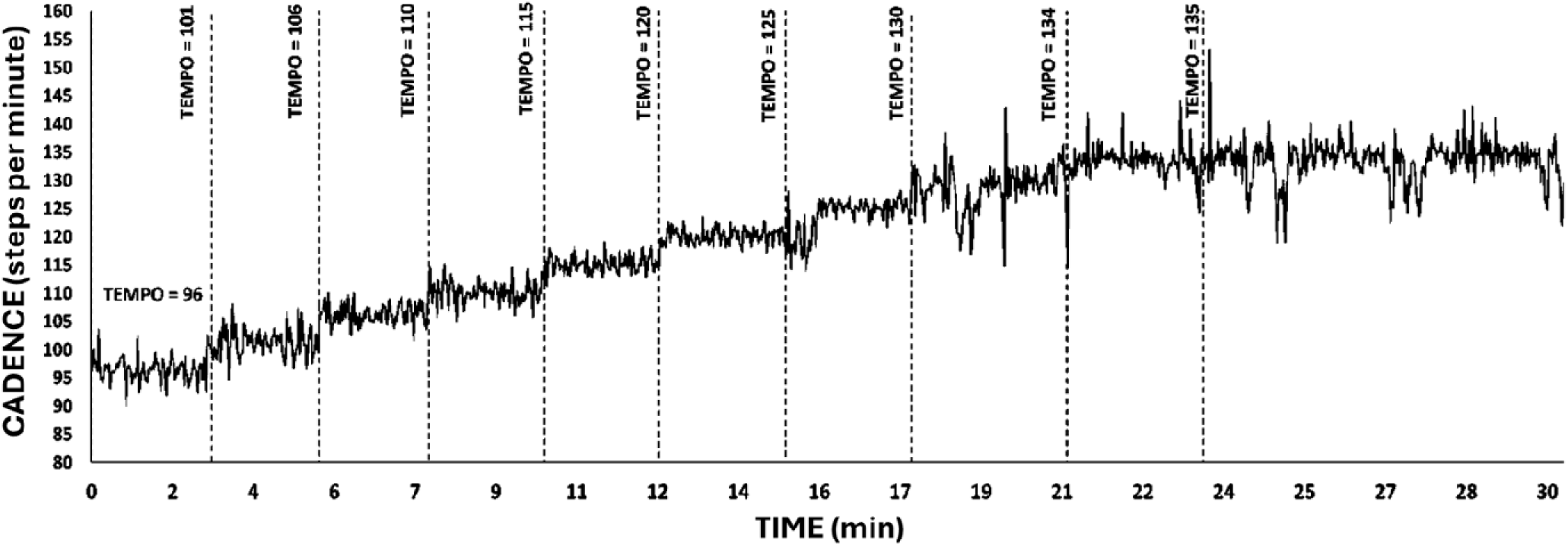
Walking cadence and target music tempo during the walking session for one participant during the RAS session.

### Knee pain

At the start of the visit, participants were trained to use the 11-point 0-10 Numeric Pain Rating Scale (NPRS) for rating their knee pain. Subsequently, participants used the NPRS to rate pain in the index and contralateral knees before the start of the RAS session, immediately at the end of the RAS session (both maximal pain during the session and current pain), and 2-3 minutes after the end of the RAS session.

### Exercise Intensity

At the start of the visit, participants were trained to use the Rating of Perceived Exertion (**RPE**, range 6-20) scale. At the end of the RAS session, participants reported the highest RPE they experienced during the session. Prior to the RAS session, participants wore a heart rate monitor on the arm (OH1+, Polar Electro Inc, Kempele, Finland) and continuous heart rate data were collected throughout the RAS session. We used the heart rate data to estimate time spent in various exercise intensity zones using maximum heart rate (**HRmax**) based on American College of Sports Medicine (ACSM) guidelines (42, 43). During the 30- minute training phase of the RAS session, we quantified the time spent in light (<64% HRmax), moderate (64-76% HRmax), and vigorous (77-95% HRmax) exercise intensity zones.

### Patient-reported Outcomes

At the end of the RAS session, participants completed the Physical Activity Enjoyment Scale (PACES) (44) to rate their experience with the device and provided feasibility feedback via a custom questionnaire. This questionnaire asked the participants to rate their comfort with the device (very uncomfortable, uncomfortable, comfortable, very comfortable), difficulty of matching their steps to the beats of the music or ability to entrain (very difficult, difficult, easy, very easy), satisfaction with the music choices (yes, no), and willingness to use the device at home (yes, no, unsure). For those who were willing to use the device at home, they were further asked about weeks of home use (< 1 week, 1 week, 2-4 weeks, 5-6 weeks, > 6 weeks), duration of each session at home (< 15 min, 15-30 min, 30-45 min, 60 min, > 60 min), and number of days per week of home use (0–7). Open-ended questions about what would make the device more engaging or any other feedback about the device were also included. Participants also completed the Knee injury and Osteoarthritis Outcome Scale (KOOS) (45).

### Analysis

Our outcomes for device feasibility were the changes in gait measures (speed, cadence, stride length) during the last 5 minutes of the RAS session compared to the baseline period as measured by the OPAL sensors. While gait measures were collected bilaterally, only data for the index leg are reported here. We also examined gait variability and symmetry as described earlier. Exercise intensity was assessed with RPE and HRmax. We assessed safety as number of adverse or serious adverse events during the RAS session. Potential device acceptability was assessed with PACES and the custom acceptability questionnaire. We also explored if the pain during the RAS session or a person’s rating of difficulty matching their steps to the beats of the music were related to the increase in cadence and gait speed during the session. For the former, we assessed Pearson’s correlation between change in pain and change in cadence/speed and for the latter, we compared the changes in cadence/speed between people who reported it being easy/very easy vs. difficult/very difficult to match their steps to the beats of the music. No sample size estimation was performed *a priori*. All data are reported descriptively and no hypothesis testing was performed.

## RESULTS

### Participant characteristics

The online pre-screening was completed by 44 persons of which 15 were enrolled (**Figure 3**). Participants were over 65 with an average BMI in the overweight range (**Table 1**). A majority self-identified as women and White and had moderate OA-related disability (KOOS scores range 40-70).

**FIGURE 3.**
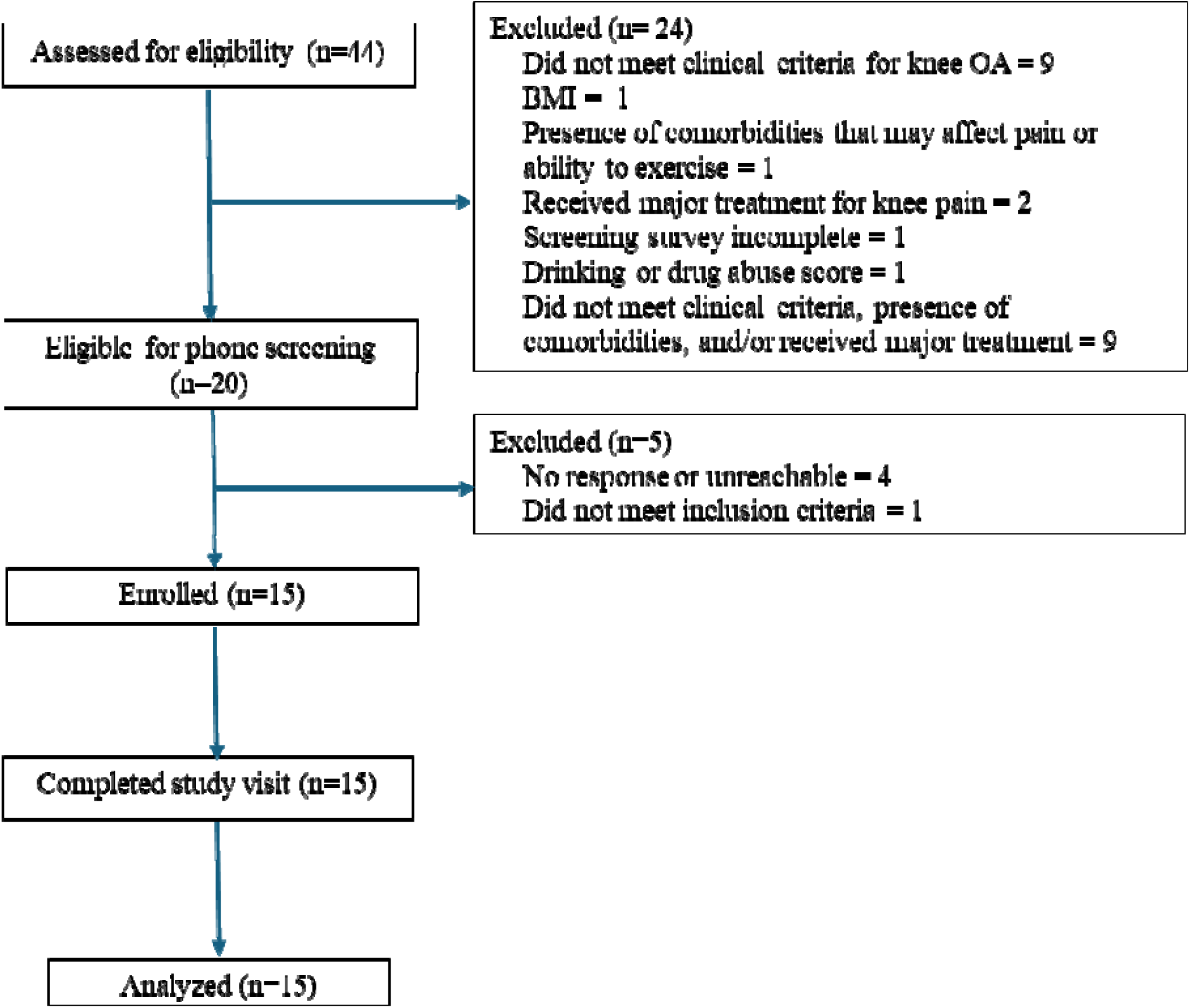
Participant flow.

**Tables 1.** Participant Characteristics. Mean and standard deviation are reported unless indicated otherwise.

| **Participants** | **n=15** |
| --- | --- |
| **Age, years** | 70.0 ± 8.6 |
| **Women, n (%)** | 10 (67%) |
| **BMI, kg/m<sup>2</sup>** | 29.4 ± 5.9 |
| **KOOS (0-100)** |  |
| *Pain* | 64.6 ± 9.6 |
| *ADL* | 70.0 ± 12.9 |
| *Symptoms* | 68.8±12.5 |
| *Sports/Recreation* | 41.3±17.0 |
| *QOL* | 52.1±12.6 |
| **Education, n (%)** |  |
| *with undergraduate degree* | 5 (33%) |
| *with graduate degree* | 9 (60%) |
| *Other\** | 1 (7%) |
| **Annual family income, n (%)** |  |
| *Between \$25,000 and \$50,000* | 4 (27%) |
| *≥\$50,000* | 8 (53%) |
| *< \$25,000, unknown, or did not report\** | 3 (20%) |
| **Race, n (%)\*** |  |
| *White* | 13 (87%) |
| *Other\** | 2 (13%) |

*\*Categories combined to maintain privacy*
*BMI = Body Mass Index; KOOS = Knee injury and Osteoarthritis Outcome Score; ADL = Activities of Daily Living; QOL = Quality of Life*

### Gait

Participants completed 30 (range 23-30) median minutes and walked on average 2455 steps (range 1916 to 2930) during the training phase of the RAS session. When comparing the last five minutes of the training to the baseline of the RAS session, participants demonstrated a 12.1% increase in cadence (Δ 12.5 [6.0, 18.9] steps per minute), a 29.5% increase in gait speed (Δ 0.3 [0.2, 0.4] m/s), and 14.8% increase in stride length (Δ 0.2 [0.1, 0.2] m) (**Figure 4**). Importantly, despite increases in gait speed, gait variability and gait symmetry did not worsen from baseline to last 5 minutes of the RAS session (**Supplementary** Figure 1). Using a threshold of 10% increase, 9/15, 11/15, and 9/15 met the threshold for cadence, gait speed, and stride length (**Figure 5**). Using a threshold of 5% increase, 10/15, 13/15, and 13/15 met the threshold for cadence, gait speed, and stride length (**Figure 5**). When comparing the carryover (with music-based RAS turned off) to the baseline, 5.2% increase in cadence, 15.9% increase in gait speed, and 9.5% increase in stride length were still present (**Figure 4**).

**FIGURE 4.**
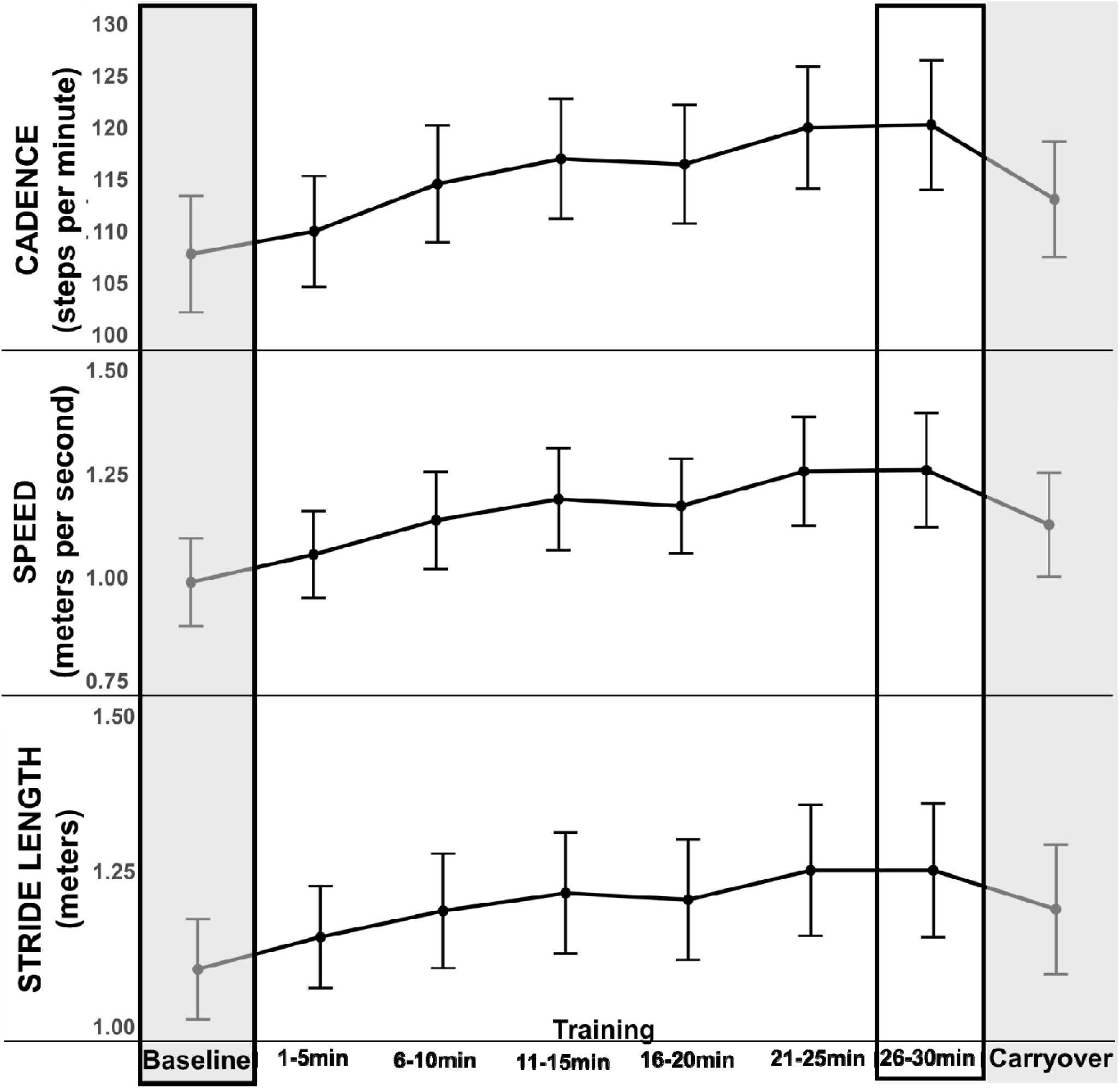
Cadence, speed, and stride length during the RAS session. The shaded vertical grey zones represent baseline and carryover phases. The black boxes indicate the time periods used to calculate the change in gait measures from baseline to last 5-minutes of training while using the device.

**FIGURE 5.**
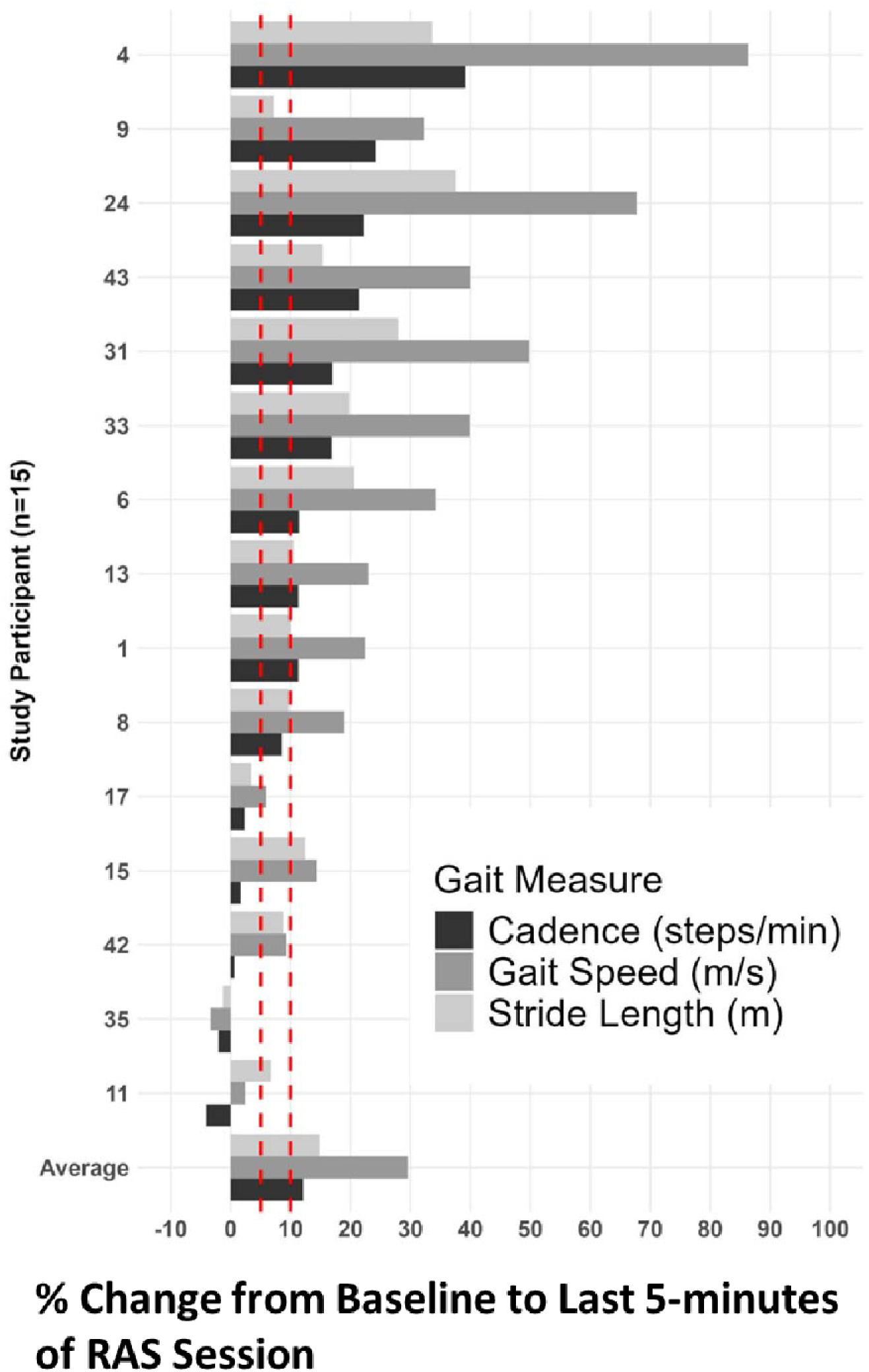
Individual changes in cadence, speed, and stride length (assessed as % change) during the last 5-minutes of the training phase vs. baseline of the RAS session. The participants are ordered by the largest to smallest change in cadence. Dotted red lines denote change of 5% and 10%.

### Exercise intensity

Maximum RPE during the RAS session was on average 13.3 (range 12-15) indicating moderate intensity of exercise. Based on HRmax, 20%, 37%, and 22% of the 30- minute training session was spent in light, moderate, and vigorous intensity exercise, respectively.

### Safety

There were no falls during the RAS session and no participants required support from the researcher; 3/15 and 1/15 participants took one and two breaks, respectively. Of these, one participant took a break due to new onset hip pain that resolved at the end of the RAS session and two participants took breaks due to fatigue. Additionally, for two participants, the RAS session had to be paused to fix issues related to OPAL or heart rate sensors. Knee pain increased by ∼ 2.4 points in the index knee during the RAS session but returned to almost pre-RAS levels within 1-2 minutes after the end of the session (**Supplementary Figure 2**).

### Acceptability

There were no technical issues with the device for any of the participants. PACES results showed that participants rated the session as enjoyable, absorbing, fun, and energizing among other attributes (mean scores >4 on 7-point scales, **Supplementary Figure 3**). All 15 participants rated the comfort with the device as comfortable or very comfortable and ∼ 73% (11/15) found it easy or very easy to match their steps to the beats of the music (i.e., entrain). For willingness to use at home, ∼87% (13/15) were willing to use the device at home and 2/15 were unsure. Of those who were willing to use the device at home, ∼77% (10/13) mentioned being willing to use it for more than 6 weeks, and the rest were willing to use it from 2-4 weeks or 5-6 weeks. Of those who were willing to use the device at home, everyone was willing to use the device for at least 30 minutes at a time; for numbers of days per week, the participants identified 4 days/week (range 2-7) as the most frequent choice. In our cohort, 80% (12/15) were satisfied the music choices available in the device and rest expressed a desire for more choices. Preferences for further improving engagement with the device included more options for musical genres, smoother transitions between songs, computability with other commercial headphones (e.g., AirPods), and a low cost.

### Factors related to changes in gait

The correlations between changes in pain and changes in cadence (r=0.11) and speed (r = -0.10) were small. People who found it easy/very easy to match their steps to the beats of the music (n=11) vs. those who found it difficult/very difficult (n=4) had ∼ 14% vs. 7% increase in cadence and ∼ 34% vs. 16% increase in gait speed during the RAS session.

## DISCUSSION

Our objective was to examine the feasibility of a wearable device to deliver a closed-loop music-based RAS intervention for people with knee OA. In this single session study, participants were safely able to increase their cadence and gait speed while walking with the device at a moderate exercise intensity under supervision. The participants rated their experience with the device positively and were willing to use the device at home. Our preliminary findings suggest that this device merits further examination as a potential intervention to improve gait quality and quantity for people with knee OA.

In our study, participants were able to increase their cadence > 10% on average while using the wearable music-based RAS device. We have previously reported associations between low cadence and increased risk of worsening pain and physical function in people with knee OA (12). Lower cadence is also related to greater knee joint loading and risk of cartilage damage independent of walking speed in people with knee OA (1, 2). Increasing cadence reduces knee loading in runners and reduces pain in individuals with patellofemoral pain (46–50). Hence, walking with higher cadence could be a promising therapeutic strategy for people with knee OA. Participants also increased their speed ∼ 30% on average while wearing the device. Lower walking speed in people with knee OA leads to increased mortality risk and hence is an important therapeutic target (51, 52). Increases in gait speed are often a secondary consequence of exercise interventions for knee OA, (53) but interventions that directly target gait speed, or those that target cadence, are largely understudied for people with knee OA. While the increases that we observed for gait speed of ∼ 0.3 m/s are larger than the reported clinically important differences ranging from 0.06-0.11 m/s for people with knee OA (54), controlled studies are needed to ascertain if these increases will translate into improvements in comfortable walking speed with longer-term use of this device. We also observed ∼ 15% increase in stride length on average. There is some evidence that longer step length is related to greater knee loading and risk of structural worsening of knee OA (55, 56). It is likely that the benefits of increases in cadence and gait speed, and engaging in moderate intensity exercise, outweigh any potential negative effects of increase in stride length. However, this will need to be confirmed in future studies.

In a 4-week feasibility study of a similar wearable closed-loop music-based RAS intervention as our study, participants with Parkinson’s disease (n=23) experienced increases in gait speed, stride length, and cadence within sessions and reductions in gait variability (57). After 4-weeks, there were improvements in moderate intensity walking, number of steps per day, quality of life, disease severity, and functional mobility. Importantly, participants completed 87% of the sessions, and no adverse events or falls were reported; participants reported that the system was easy to use. A single-session feasibility study with a similar intervention reported increases in gait speed and cadence in people with stroke (n=11) (58). More recently a multi-site parallel-arm efficacy randomized controlled trial of a wearable closed-loop music-based RAS vs. active control was completed in people with stroke (n=87) (59). Results showed greater improvements in gait speed and greater proportion of treatment responders in the intervention arm. Intervention was determined to be safe. Based on the results of this study, a closed-loop music-based RAS intervention is currently commercially available in the US by prescription for adults with chronic stroke with walking impairment. These prior studies and our findings from this device feasibility study suggest that this wearable closed-loop music-based RAS device can be a potential novel approach to improve gait quality and quantity in people with knee OA.

There was some variability across participants in terms of changes in gait while walking with the device. Of the 15, five did not show at least a 5% increase in cadence while wearing the device. However, only two did not show at least a 5% increase in gait speed, suggesting that at least a few participants achieved increases in gait speed by increasing their stride length alone. The correlations between changes in pain and gait measures were small and unlikely to be clinically meaningful. Hence, knee pain does not appear to interfere with a person’s ability to improve their gait quality while using the RAS device. Although our sample size is quite small, our exploratory results show that people who found it easy/very easy to match their steps to the beats of the music had almost double the amount of increase in cadence and gait speed. There is some prior evidence to support the speculation that beat perception ability can influence gait parameters while walking with RAS (60–62). Importantly, auditory-motor entrainment is defined as an involuntary process and asking participants about their comfort with matching their steps to the music may not truly reflect the ability of their neuromotor system to achieve entrainment. Nonetheless, whether a person’s ability to perceive the beats and/or entrain influences their response to a longer-term intervention with this device should be examined in future studies.

While walking with the device, all participants reported an RPE between 12-15 reflecting a moderate intensity exertion. These self-reported data were consistent with the objective findings from the heart rate measurements with participants spending ∼ 60% of the time in moderate-to-vigorous intensity exercise zone based on HRmax. Our findings are in line with studies showing that increasing cadence can be a strategy to achieve moderate intensity effort during walking exercise indicated by %HRmax of 64-76% and RPE of 12-13 (13)(63). It needs to be examined whether people with knee OA can use this device for sustained engagement in physical activity while meeting the 150-300 minutes per week of moderate intensity exercise guidelines.

In this single supervised session, the device was found to be safe. There were no serious adverse events (e.g., falls). Only one participant experienced new onset pain in the hip that was resolved at the end of the session. While there were small increases in knee pain while walking with the device as expected for people with knee OA, the pain largely subsided to pre-RAS levels within a few minutes after the end of the session. We have previously reported that small increase in pain with exercise is related to greater likelihood of experiencing exercise-induced hypoalgesia (64). Two participants took breaks due to fatigue; exercise tolerance would be expected to improve over the course of a longer intervention. For a longer intervention with this device where the participants are expected to use the device on their own, safety monitoring would be an important aspect including screening for fall risks, any cardiac or other conditions that may preclude a person from engaging in moderate intensity exercise and educating the participants about acceptable and unacceptable levels of pain.

Participants in our study were enthusiastic about the device. They reported a largely positive experience across several enjoyment domains assessed in PACES. Some participants were not satisfied with the music choices, indicating the potential for further refinement. A majority of the participants were willing to use the device at home, at least 30 minutes per session, for 4 days per week, for at least 6 weeks. These findings provide further support for examining the potential of this device as digital therapeutic strategy for people with knee OA.

Our findings need to be interpreted considering study limitations. This was a device feasibility study and not a study of the effects of this intervention on health-related outcomes. It is important to acknowledge that the reported changes in gait while walking with the device are likely not reflective of potential therapeutic effects of this device if used over the long-term. We did not select our participants to be physically inactive. Also, our cohort was not racially diverse, with a majority having at least an undergraduate degree and annual family income > $50,000. Hence, our findings may not be generalizable to people with knee OA who differ from our cohort in terms of their physical activity level, racial background, education or income levels, BMI, and levels of disability.

To conclude, this single-session feasibility study of a wearable closed-loop music-based RAS device demonstrates increases in cadence and gait speed while safely walking at a moderate exercise intensity under supervision in people with knee OA. Participants reported a positive experience with the device and were willing to use the device at home for longer durations. Longer-term randomized studies to examine whether this device can improve both quality and quantity of walking for people with knee OA are needed.

## Data Availability

All data produced in the present study are available upon reasonable request to the authors

## ACKNOWLEDGMENTS

The authors would like to thank the participants of this study.

## AUTHOR CONTRIBUTIONS

Dr. Kumar had full access to all of the data in the study and took responsibility for the integrity of the data and the accuracy of the data analysis. Study conception and design: Kumar Acquisition of data: Yang, Okamura, Kim Analysis and interpretation of data: All authors Manuscript drafting and approval: All authors

## SUPPLEMENTRAY MATERIAL

**SUPPLEMENTARY FIGURE 1.**
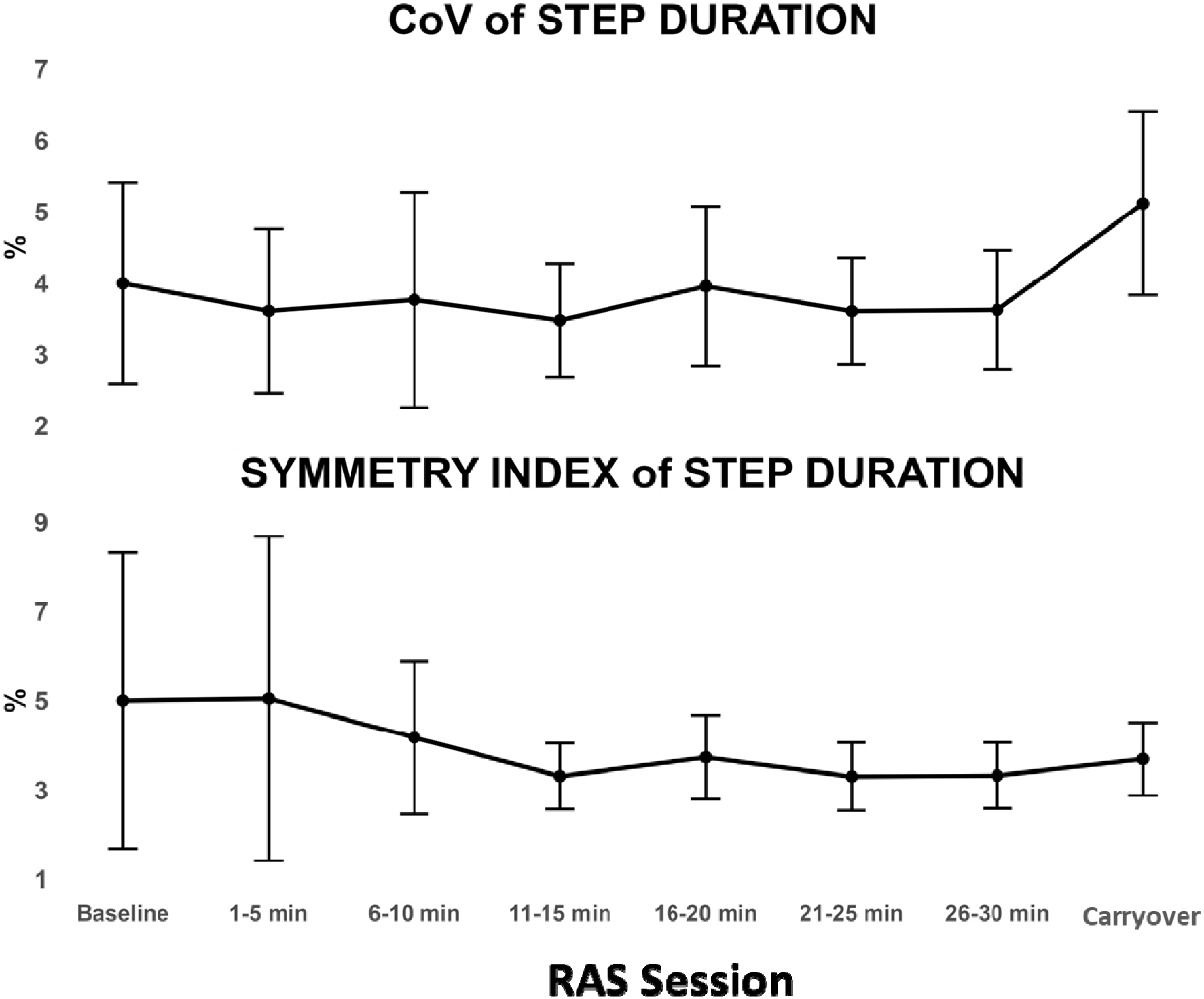
Coefficient of Variation (CoV) of step duration (TOP) and symmetry index of step duration (BOTTOM) during the RAS session.

**SUPPLEMENTARY FIGURE 2:**
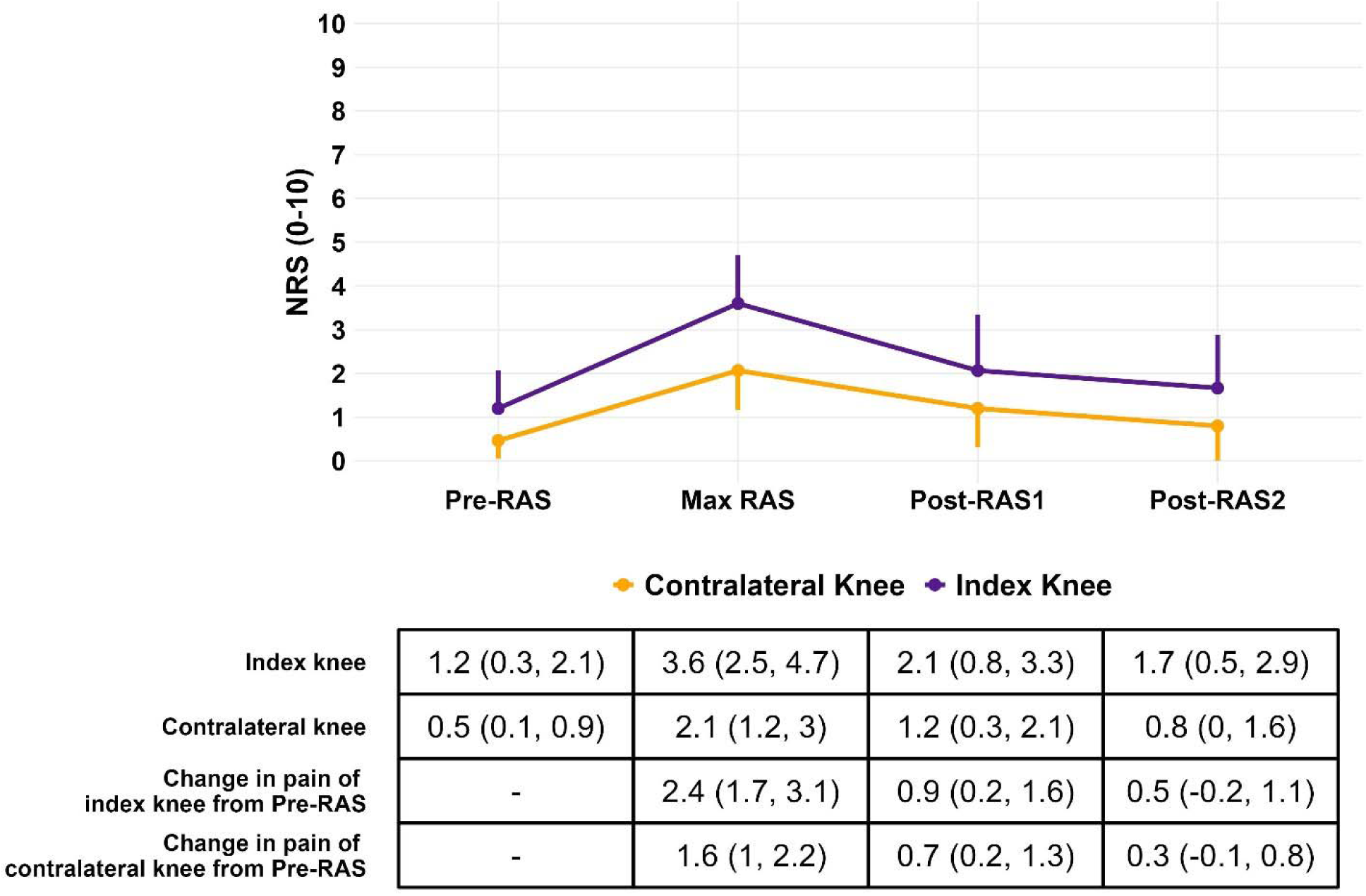
Pain in the index and contralateral knee on the 0-10 numeric pain rating scale during the visit. All data are reported as mean (95% confidence intervals). Pre-RAS refers to before the start of the RAS session; Max RAS refers to pain rating of maximum knee pain during the RAS session; Post-RAS1 refers to pain rating immediately at the end of the RAS session; Post-RAS2 refers to pain rating 2-3 minutes after the end of the RAS session.

**SUPPLEMENTARY FIGURE 3.**
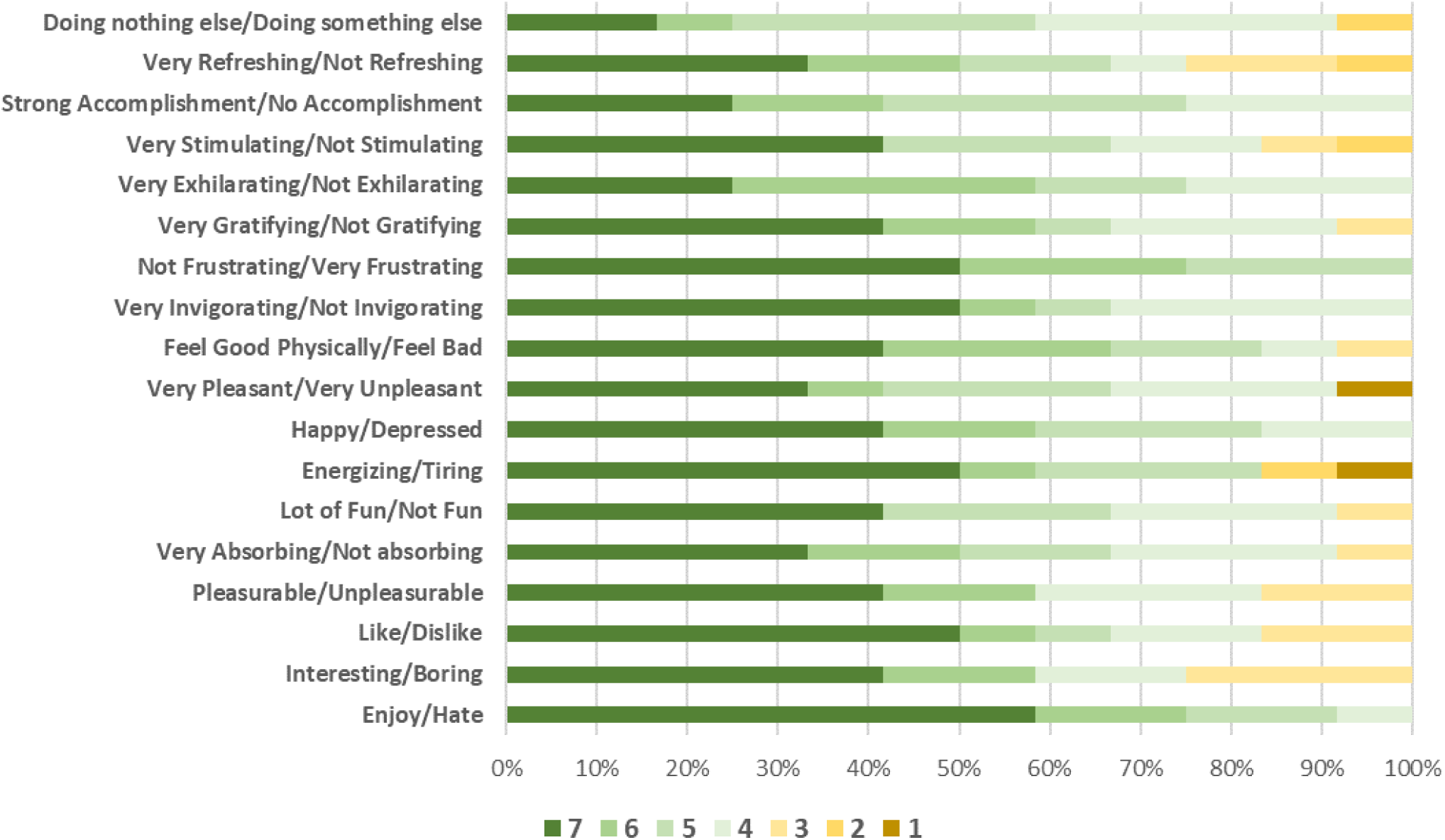
Proportion of participants in each response category (range 1-7) for each item of the Physical Activity Enjoyment Scale (PACES). Where necessary, item responses have been reordered such that higher scores indicate a more positive response.

## REFERENCES

1. Hart HF, Birmingham TB, Primeau CA, Pinto R, Leitch K, Giffin JR. Associations Between Cadence and Knee Loading in Patients With Knee Osteoarthritis. Arthritis care & research. 2021;73(11):1667–71.

2. Hart HF, Gross KD, Crossley KM, Barton CJ, Felson DT, Guermazi A, et al. Step Rate and Worsening of Patellofemoral and Tibiofemoral Joint Osteoarthritis in Women and Men: The Multicenter Osteoarthritis Study. Arthritis care & research. 2020;72(1):107–13.

3. Miyazaki T, Wada M, Kawahara H, Sato M, Baba H, Shimada S. Dynamic load at baseline can predict radiographic disease progression in medial compartment knee osteoarthritis. AnnRheumDis. 2002;61(7):617–22.

4. Brown C, Marinko L, LaValley MP, Kumar D. Quadriceps Strength After Anterior Cruciate Ligament Reconstruction Compared With Uninjured Matched Controls: A Systematic Review and Meta-analysis. Orthop J Sports Med. 2021;9(4):2325967121991534.

5. Kumar D, Manal KT, Rudolph KS. Knee joint loading during gait in healthy controls and individuals with knee osteoarthritis. Osteoarthritis and cartilage. 2013;21(2):298–305.

6. Bennell KL, Bowles KA, Wang Y, Cicuttini F, Davies-Tuck M, Hinman RS. Higher dynamic medial knee load predicts greater cartilage loss over 12 months in medial knee osteoarthritis. Ann Rheum Dis. 2011;70(10):1770–4.

7. Hodges PW, van den Hoorn W, Wrigley TV, Hinman RS, Bowles KA, Cicuttini F, et al. Increased duration of co-contraction of medial knee muscles is associated with greater progression of knee osteoarthritis. Manual therapy. 2016;21:151–8.

8. Wink AE, Gross KD, Brown CA, Guermazi A, Roemer F, Niu J, et al. Varus thrust during walking and the risk of incident and worsening medial tibiofemoral MRI lesions: the Multicenter Osteoarthritis Study. Osteoarthritis and cartilage. 2017;25(6):839–45.

9. Chang AH, Song J, Lee J, Chang RW, Semanik PA, Dunlop DD. Proportion and associated factors of meeting the 2018 Physical Activity Guidelines for Americans in adults with or at risk for knee osteoarthritis. Osteoarthritis and cartilage. 2020;28(6):774–81.

10. Kolasinski SL, Neogi T, Hochberg MC, Oatis C, Guyatt G, Block J, et al. 2019 American College of Rheumatology/Arthritis Foundation Guideline for the Management of Osteoarthritis of the Hand, Hip, and Knee. Arthritis Rheumatol. 2020;72(2):220–33.

11. Bannuru RR, Osani MC, Vaysbrot EE, Arden NK, Bennell K, Bierma-Zeinstra SMA, et al. OARSI guidelines for the non-surgical management of knee, hip, and polyarticular osteoarthritis. Osteoarthritis and cartilage. 2019.

12. Bacon KL, Felson DT, Jafarzadeh SR, Kolachalama VB, Hausdorff JM, Gazit E, et al. Gait Alterations and Association With Worsening Knee Pain and Physical Function: A Machine Learning Approach With Wearable Sensors in the Multicenter Osteoarthritis Study. Arthritis care & research. 2024;76(7):984–92.

13. Slaght J, Sénéchal M, Hrubeniuk TJ, Mayo A, Bouchard DR. Walking Cadence to Exercise at Moderate Intensity for Adults: A Systematic Review. J Sports Med (Hindawi Publ Corp). 2017;2017:4641203.

14. Merchant H, Grahn J, Trainor L, Rohrmeier M, Fitch WT. Finding the beat: a neural perspective across humans and non-human primates. Philos Trans R Soc Lond B Biol Sci. 2015;370(1664):20140093.

15. Magee WL, Clark I, Tamplin J, Bradt J. Music interventions for acquired brain injury. Cochrane Database Syst Rev. 2017;1(1):Cd006787.

16. Nascimento LR, de Oliveira CQ, Ada L, Michaelsen SM, Teixeira-Salmela LF. Walking training with cueing of cadence improves walking speed and stride length after stroke more than walking training alone: a systematic review. J Physiother. 2015;61(1):10–5.

17. Ghai S, Ghai I, Schmitz G, Effenberg AO. Effect of rhythmic auditory cueing on parkinsonian gait: A systematic review and meta-analysis. Sci Rep. 2018;8(1):506.

18. Shahraki M, Sohrabi M, Taheri Torbati HR, Nikkhah K, NaeimiKia M. Effect of rhythmic auditory stimulation on gait kinematic parameters of patients with multiple sclerosis. J Med Life. 2017;10(1):33–7.

19. Chauvigné LA, Gitau KM, Brown S. The neural basis of audiomotor entrainment: an ALE meta-analysis. Front Hum Neurosci. 2014;8:776.

20. Thaut MH. The discovery of human auditory-motor entrainment and its role in the development of neurologic music therapy. Prog Brain Res. 2015;217:253–66.

21. Ye X, Li L, He R, Jia Y, Poon W. Rhythmic auditory stimulation promotes gait recovery in Parkinson’s patients: A systematic review and meta-analysis. Front Neurol. 2022;13:940419.

22. Ghai S, Ghai I, Schmitz G, Effenberg AO. Effect of rhythmic auditory cueing on parkinsonian gait: A systematic review and meta-analysis. Scientific Reports. 2018;8(1):506.

23. Hurt CP, Rice RR, McIntosh GC, Thaut MH. Rhythmic Auditory Stimulation in Gait Training for Patients with Traumatic Brain Injury. J Music Ther. 1998;35(4):228–41.

24. Ghai S, Ghai I. Effects of Rhythmic Auditory Cueing in Gait Rehabilitation for Multiple Sclerosis: A Mini Systematic Review and Meta-Analysis. Front Neurol. 2018;9:386.

25. Ghai S, Ghai I, Effenberg AO. Effect of rhythmic auditory cueing on gait in cerebral palsy: a systematic review and meta-analysis. Neuropsychiatr Dis Treat. 2018;14:43–59.

26. Bella SD, Benoit CE, Farrugia N, Schwartze M, Kotz SA. Effects of musically cued gait training in Parkinson’s disease: beyond a motor benefit. Ann N Y Acad Sci. 2015;1337:77–85.

27. Zatorre RJ, Chen JL, Penhune VB. When the brain plays music: auditory-motor interactions in music perception and production. Nat Rev Neurosci. 2007;8(7):547–58.

28. Ottaviani S, Bernard JL, Bardin T, Richette P. Effect of music on anxiety and pain during joint lavage for knee osteoarthritis. Clin Rheumatol. 2012;31(3):531–4.

29. Hsu CC, Chen SR, Lee PH, Lin PC. The Effect of Music Listening on Pain, Heart Rate Variability, and Range of Motion in Older Adults After Total Knee Replacement. Clin Nurs Res. 2019;28(5):529–47.

30. Innes KE, Selfe TK, Kandati S, Wen S, Huysmans Z. Effects of Mantra Meditation versus Music Listening on Knee Pain, Function, and Related Outcomes in Older Adults with Knee Osteoarthritis: An Exploratory Randomized Clinical Trial (RCT). Evid Based Complement Alternat Med. 2018;2018:7683897.

31. Cheever T, Taylor A, Finkelstein R, Edwards E, Thomas L, Bradt J, et al. NIH/Kennedy Center Workshop on Music and the Brain: Finding Harmony. Neuron. 2018;97(6):1214–8.

32. Brown EL, Gannotti ME, Veneri DA. Including Arts in Rehabilitation Enhances Outcomes in the Psychomotor, Cognitive, and Affective Domains: A Scoping Review. Phys Ther. 2022;102(4).

33. Hsu HF, Chen KM, Belcastro F. The effect of music interventions on chronic pain experienced by older adults: A systematic review. J Nurs Scholarsh. 2022;54(1):64–71.

34. Sihvonen AJ, Pitkäniemi A, Särkämö T, Soinila S. Isn’t There Room for Music in Chronic Pain Management? J Pain. 2022;23(7):1143–50.

35. Usui C, Kirino E, Tanaka S, Inami R, Nishioka K, Hatta K, et al. Music Intervention Reduces Persistent Fibromyalgia Pain and Alters Functional Connectivity Between the Insula and Default Mode Network. Pain Med. 2020;21(8):1546–52.

36. Lee JH. The Effects of Music on Pain: A Meta-Analysis. J Music Ther. 2016;53(4):430–77.

37. Loewy J. Underlying Music Mechanisms Influencing the Neurology of Pain: An Integrative Model. Brain Sci. 2022;12(10).

38. Park KS, Williams DM, Etnier JL. Exploring the use of music to promote physical activity: From the viewpoint of psychological hedonism. Front Psychol. 2023;14:1021825.

39. Washabaugh EP, Kalyanaraman T, Adamczyk PG, Claflin ES, Krishnan C. Validity and repeatability of inertial measurement units for measuring gait parameters. Gait Posture. 2017;55:87–93.

40. Salarian A, Russmann H, Vingerhoets FJ, Dehollain C, Blanc Y, Burkhard PR, et al. Gait assessment in Parkinson’s disease: toward an ambulatory system for long-term monitoring. IEEE Trans Biomed Eng. 2004;51(8):1434–43.

41. Robinson RO, Herzog W, Nigg BM. Use of force platform variables to quantify the effects of chiropractic manipulation on gait symmetry. J Manipulative Physiol Ther. 1987;10(4):172–6.

42. Gellish RL, Goslin BR, Olson RE, Mcdonald A, Russi GD, Moudgil VK. Longitudinal Modeling of the Relationship between Age and Maximal Heart Rate. Medicine & Science in Sports & Exercise. 2007;39(5):822–9.

43. Schulz JM, Birmingham TB, Atkinson HF, Woehrle E, Primeau CA, Lukacs MJ, et al. Are we missing the target? Are we aiming too low? What are the aerobic exercise prescriptions and their effects on markers of cardiovascular health and systemic inflammation in patients with knee osteoarthritis? A systematic review and meta-analysis. Br J Sports Med. 2020;54(13):771–5.

44. Kendzierski D, DeCarlo KJ. Physical Activity Enjoyment Scale: Two Validation Studies. Journal of Sport and Exercise Psychology. 1991;13(1):50–64.

45. Roos EM, Roos HP, Lohmander LS, Ekdahl C, Beynnon BD. Knee Injury and Osteoarthritis Outcome Score (KOOS)--development of a self-administered outcome measure. J Orthop Sports Phys Ther. 1998;28(2):88–96.

46. Heiderscheit BC, Chumanov ES, Michalski MP, Wille CM, Ryan MB. Effects of step rate manipulation on joint mechanics during running. Medicine and science in sports and exercise. 2011;43(2):296–302.

47. Willy RW, Willson JD, Clowers K, Baggaley M, Murray N. The effects of body-borne loads and cadence manipulation on patellofemoral and tibiofemoral joint kinetics during running. J Biomech. 2016;49(16):4028–33.

48. Bramah C, Preece SJ, Gill N, Herrington L. A 10% Increase in Step Rate Improves Running Kinematics and Clinical Outcomes in Runners With Patellofemoral Pain at 4 Weeks and 3 Months. Am J Sports Med. 2019;47(14):3406–13.

49. Neal BS, Barton CJ, Birn-Jeffrey A, Daley M, Morrissey D. The effects & mechanisms of increasing running step rate: A feasibility study in a mixed-sex group of runners with patellofemoral pain. Phys Ther Sport. 2018;32:244–51.

50. Esculier JF, Bouyer LJ, Roy JS. Running gait modifications can lead to immediate reductions in patellofemoral pain. Front Sports Act Living. 2022;4:1048655.

51. Master H, Neogi T, Callahan LF, Nelson AE, LaValley M, Cleveland RJ, et al. The association between walking speed from short- and standard-distance tests with the risk of all-cause mortality among adults with radiographic knee osteoarthritis: data from three large United States cohort studies. Osteoarthritis Cartilage. 2020;28(12):1551–8.

52. Master H, Neogi T, LaValley M, Thoma LM, Zhang Y, Voinier D, et al. Does the 1-year Decline in Walking Speed Predict Mortality Risk Beyond Current Walking Speed in Adults With Knee Osteoarthritis? J Rheumatol. 2021;48(2):279–85.

53. Li S, Ng WH, Abujaber S, Shaharudin S. Effects of resistance training on gait velocity and knee adduction moment in knee osteoarthritis patients: a systematic review and meta-analysis. Scientific Reports. 2021;11(1):16104.

54. Gilbert AL, Song J, Cella D, Chang RW, Dunlop DD. What Is an Important Difference in Gait Speed in Adults With Knee Osteoarthritis? Arthritis Care Res (Hoboken). 2021;73(4):559–65.

55. Stefanik JJ, Gross KD, Guermazi A, Felson DT, Roemer FW, Niu J, et al. Relation of Step Length to Magnetic Resonance Imaging-Detected Structural Damage in the Patellofemoral Joint: The Multicenter Osteoarthritis Study. Arthritis Care Res (Hoboken). 2016;68(6):776–83.

56. Bacon K, Felson D, Jafarzadeh SR, Hausdorff JM, Gazit E, Segal N, et al., editors. Association of wearable-sensor derived gait measures with cartilage worsening over 2 years in the Multicenter Osteoarthritis Study. OARSI Annual Congress; 2024; Vienna, Austria: Osteoarthritis and Cartilage.

57. Zajac JA, Porciuncula F, Cavanaugh JT, McGregor C, Harris BA, Smayda KE, et al. Feasibility and Proof-of-Concept of Delivering an Autonomous Music-Based Digital Walking Intervention to Persons with Parkinson’s Disease in a Naturalistic Setting. J Parkinsons Dis. 2023.

58. Hutchinson K, Sloutsky R, Collimore A, Adams B, Harris B, Ellis TD, et al. A Music-Based Digital Therapeutic: Proof-of-Concept Automation of a Progressive and Individualized Rhythm-Based Walking Training Program After Stroke. Neurorehabil Neural Repair. 2020;34(11):986–96.

59. Awad LN, Jayaraman A, Nolan KJ, Lewek MD, Bonato P, Newman M, et al. Efficacy and safety of using auditory-motor entrainment to improve walking after stroke: a multi-site randomized controlled trial of InTandemTM. Nature Communications. 2024;15(1):1081.

60. Ready EA, McGarry LM, Rinchon C, Holmes JD, Grahn JA. Beat perception ability and instructions to synchronize influence gait when walking to music-based auditory cues. Gait Posture. 2019;68:555–61.

61. Roberts BS, Ready EA, Grahn JA. Musical enjoyment does not enhance walking speed in healthy adults during music-based auditory cueing. Gait & Posture. 2021;89:132–8.

62. Ready EA, Holmes JD, Grahn JA. Gait in younger and older adults during rhythmic auditory stimulation is influenced by groove, familiarity, beat perception, and synchronization demands. Hum Mov Sci. 2022;84:102972.

63. McAvoy CR, Moore CC, Aguiar EJ, Ducharme SW, Schuna JM, Barreira TV, et al. Cadence (steps/min) and relative intensity in 21 to 60-year-olds: the CADENCE-adults study. International Journal of Behavioral Nutrition and Physical Activity. 2021;18(1):27.

64. Lee S, Neogi T, Senderling B, Gheller M, Adamowicz L, Georgiev P, et al., editors. No Pain, No Gain: Association Of Pain Increase During Exercise With Exercise-Induced Hypoalgesia In People With Knee Osteoarthriti. OARSI Annual Congress; 2023; Denver, CO.

